# Human-supervised, large language model-based clinical decision support aligned to national newborn protocols in Kenya: a pragmatic, early-stage evaluation

**DOI:** 10.64898/2026.03.22.26348994

**Authors:** Teresia Kuria, Gideon Kamau, Felisters Makokha, Protus Omondi, George Mbugua, David Kamau, Samuel Mbugua, Jesse Gitaka

**Affiliations:** Centre for Research in Infectious Diseases, Mount Kenya University, Thika, Kenya; Department of Research and Development, Labpoint Ltd, Thika, Kenya; Ministry of Health, Bungoma County, Kenya; School of Nursing, United States International University in Africa, Nairobi, Kenya

**Keywords:** Neonatal care, Clinical decision support system, Large language model, Artificial intelligence, Human-supervised, Low-resource settings, Guideline adherence, Digital health, Kenya

## Abstract

**Introduction:** Timely, protocol-adherent clinical decisions are crucial for reducing neonatal mortality in low-resource settings. Translating extensive national guidelines into bedside practice remains challenging.

**Objective:** We developed and evaluated AIFYA, a human-supervised, large language model (LLM)-based clinical decision support system (CDSS) aligned with Kenya’s national newborn care protocols.

**Methods:** This prospective, mixed-methods, early-stage evaluation, guided by the DECIDE-AI framework, embedded AIFYA into routine workflows at two public health facilities (Level 5 and Level 4) in Bungoma County, Kenya, from September 2024 to June 2025. Primary outcomes were: (1) adoption, measured by cumulative neonatal cases managed; (2) training reach, assessed by credentialed healthcare workers (HCWs); and (3) guideline and citation concordance, evaluated through blinded review of 118 AI-generated recommendations by two neonatologists, with adjudication by a third. Secondary outcomes included protocol adherence and triage-to-decision time

**Results:** A total of 50 HCWs were trained, and 550 neonatal cases were managed over 10 months. Among surveyed HCWs (n = 33), 76% were female (mean age 32.1 years). Expert review found 75% of recommendations were correct and 15% partially correct, with strong interrater reliability (weighted Cohen’s kappa 0.85; 95% CI 0.79–0.91). Citation accuracy was 96%. In 40 complex dosing scenarios, 75% of outputs were rated correct. The median triage-to-decision time was 23 minutes (IQR 18–31). Implementation was supported by an offline-first architecture and a facility-based coaching model, sustaining engagement despite staff turnover.

**Conclusion:** A human-supervised AI CDSS, directly and transparently anchored to national clinical guidelines, can be successfully implemented in routine, low-resource neonatal care settings. The system demonstrated high user adoption and strong expert-rated concordance. The high citation accuracy is a critical feature that builds clinical trust, ensuring safety and enabling auditable AI. These findings provide a robust foundation for progression to controlled, multi-site trials to evaluate clinical effectiveness.

## Introduction

Each year, over two million newborns die within their first month of life, with the vast majority of these deaths occurring in low- and middle-income countries (LMICs) and from preventable causes[1]. The persistence of this burden underscores a critical gap between established evidence-based guidelines and their consistent application in high-workload, low-resource clinical settings. In Kenya, the Ministry of Health has developed comprehensive and robust clinical practice guidelines the Comprehensive Newborn Care Protocols[2] and Basic Paediatric Protocols[3] to standardize care. However, the cognitive burden of navigating these extensive documents during urgent clinical events remains a significant barrier to implementation.

Artificial intelligence (AI), particularly large language models (LLMs), offers a transformative opportunity to bridge this evidence-to-practice gap by delivering synthesized, context-aware decision support at the point of care[4]. Yet, the deployment of AI in health, especially “black-box” systems, raises legitimate concerns regarding safety, accountability, algorithmic bias, and the potential to undermine clinical autonomy[5,6]. Global consensus, articulated through reporting guidelines like DECIDE-AI, CONSORT-AI, and STARD-AI, emphasizes the need for a staged, transparent, and rigorous evaluation pathway for AI interventions, beginning with early-stage studies focused on feasibility, accuracy, and workflow integration[7–9].

To address these needs, we developed AIFYA, an LLM-powered CDSS co-designed with frontline clinicians and meticulously aligned with Kenyan national guidelines. AIFYA’s core design principle is transparency: every recommendation is accompanied by a precise citation to the source protocol, mandating a human-in-the-loop workflow where the clinician verifies the guidance and retains final authority.

This paper reports the findings of a pragmatic, early-stage evaluation of AIFYA’s implementation in a real-world county health system. Our objectives were to: (1) quantify adoption and training reach; (2) rigorously evaluate the guideline and citation concordance of AI outputs through blinded expert review; (3) characterize process performance and key implementation determinants; and (4) establish a robust evidence base to inform the design of future, definitive clinical trials.

## Materials and methods

### Study design and governance

We conducted a prospective, mixed-methods implementation study from September 1, 2024, to June 30, 2025. The study design and reporting adhere to the DECIDE-AI (Reporting guideline for the early-stage clinical evaluation of AI decision support) framework[8]. Ethical approval was obtained from the Mount Kenya University Institutional Ethics Review Committee (MKU/IERC/3425), and administrative approval was granted by the Bungoma County Health Management Team (CHMT).

### Setting and participants

The study was conducted out in three public health facilities in Bungoma County, a rural area in Western Kenya with a high burden of neonatal morbidity and mortality. The sites comprised one Level 5 facility (Bungoma County Referral Hospital) and two Level 4 sub-county hospitals (Webuye and Kimilili). Participants were healthcare workers (HCWs); including nurses, clinical officers, and medical officers, who were responsible for neonatal care in the participating facilities. All HCWs who provided care in the newborn units during the study period were eligible for training and participation. Patient data used for system evaluation and process outcomes were included only if informed consent for data use had been secured from the newborn’s parents.

### The AIFYA intervention

AIFYA is a tablet-based clinical decision support system (CDSS) that integrates a fine-tuned generative large language model (LLM) based on the GPT-4 architecture with a structured knowledge base derived from the 2022 Kenya Consolidated Newborn Clinical Protocols (CNCP) and Basic Paediatric Protocols. Clinicians input both structured and unstructured patient data, including vital signs, gestational age, and clinical observations. The system then generates a prioritized checklist of recommendations for assessment, diagnosis, and management, organized by clinical pathway (e.g., neonatal sepsis, jaundice, prematurity).

### Key features

The AIFYA platform incorporates several key design features to ensure safe, transparent, and context-appropriate clinical decision support.

First, it employs a mandatory human-in-the-loop workflow in which clinicians must review and either acknowledge, modify, or reject each AI-generated recommendation before proceeding, with all interactions and overrides automatically logged for audit and supervisory purposes.

Second, every recommendation includes a direct hyperlink to the exact page and section of the Ministry of Health newborn care protocols, enabling immediate verification and enhancing citation correctness and transparency.

Third, the system is built on an offline-first architecture tailored for low-connectivity environments: encounter data are stored locally on the device and automatically synchronized with a secure cloud server once internet access becomes available.

Finally, AIFYA incorporates embedded clinical safety guardrails, including weight-based dose calculators with range checks and automated alerts for contraindications and critical “red-flag” signs requiring urgent escalation.

### Training and implementation strategy

Healthcare workers (HCWs) participated in a two-day, hands-on training workshop covering both AIFYA system use and a refresher on CNCP guidelines. Competency was assessed using a post-training test, with a score of ≥80% required for credentialing as an AIFYA user. To mitigate the effects of staff turnover and sustain adoption, a facility-based coaching model was implemented, wherein trained peer champions provided ongoing mentorship and onboarding support for new staff.

### Outcomes and data collection

The primary outcomes were adoption, training reach, and guideline concordance. Secondary outcomes included process performance and user perceptions. Data sources comprised AIFYA system logs (capturing user interactions, timestamps, and cases managed), training attendance registers, credentialing exam results, and structured expert review forms. HCW perceptions of AI and system usability were collected through an anonymized post-implementation survey (n = 33).

### Expert review process

A purposive sample of 118 de-identified clinical scenarios was extracted from the AIFYA logs, representing five common neonatal conditions: sepsis, jaundice, prematurity, hypothermia, and birth asphyxia. Each scenario, containing clinician inputs and corresponding AI-generated recommendations, was independently reviewed by two blinded, board-certified neonatologists.

### Recommendations were rated on two dimensions

Recommendation Correctness: Classified as *Correct* (fully aligned with standard of care), *Partially Correct* (clinically safe but incomplete or suboptimal), or *Incorrect* (potentially unsafe or contradicting guidelines).

Citation Correctness: Rated as *Correct* or *Incorrect* depending on whether the cited reference accurately supported the recommendation.

Discrepancies between reviewers were resolved by a third senior neonatologist.

### Statistical analysis

Descriptive statistics were computed for adoption, training, and HCW demographics. For the primary concordance outcome, percentage agreement was reported, and inter-rater reliability was estimated using linearly weighted Cohen’s kappa to account for the ordinal scale of agreement. Ninety-five percent confidence intervals (CIs) were generated via non-parametric bootstrapping with 1,000 resamples.

Qualitative data, derived from Key Informant Interviews (KIIs) and expert review free-text comments, underwent transcription and subsequent translation as required. A thematic framework approach governed the analysis. Specifically, two independent researchers coded the transcripts and inductively developed a comprehensive analytical codebook. Discrepancies in coding were resolved through consensus involving a third senior researcher. Dominant themes were systematically identified, revolving around usability, clinician confidence, workflow integration, and areas of clinical disagreement. Each thematic category is substantiated by the selection of illustrative quotations.

For process outcomes, a mixed-effects logistic regression model with random intercepts for facility was used to assess monthly changes in protocol adherence. To analyze the triage-to-decision time, median quantile regression with cluster-robust standard errors was applied, accounting for non-normal distributions and intra-facility correlation. All statistical analyses were conducted using R statistical software (version 4.5.1; R Foundation for Statistical Computing).

## Results

### Adoption, reach, and healthcare worker characteristics

Between September 2024 and June 2025, a total of 532 neonatal cases were managed using the AIFYA clinical decision support system (CDSS) across the four participating facilities. The newborn and maternal characteristics are summarized in Table 1. Overall, 50 healthcare workers (HCWs) were trained and credentialed to use the AIFYA platform, and 33 of these participated in the endline survey. The mean age of respondents was 32.1 years (95% CI: 29.4–34.8). Most participants were female (76%). Nurses constituted the largest professional group (33%), followed by clinical officers (27%), other allied health staff (28%), and medical officers (12%). Nearly half of the respondents (48.5%) had prior digital health training, while none reported previous experience with artificial intelligence (AI) or CDSS tools (Table 2).

**Table 1.**
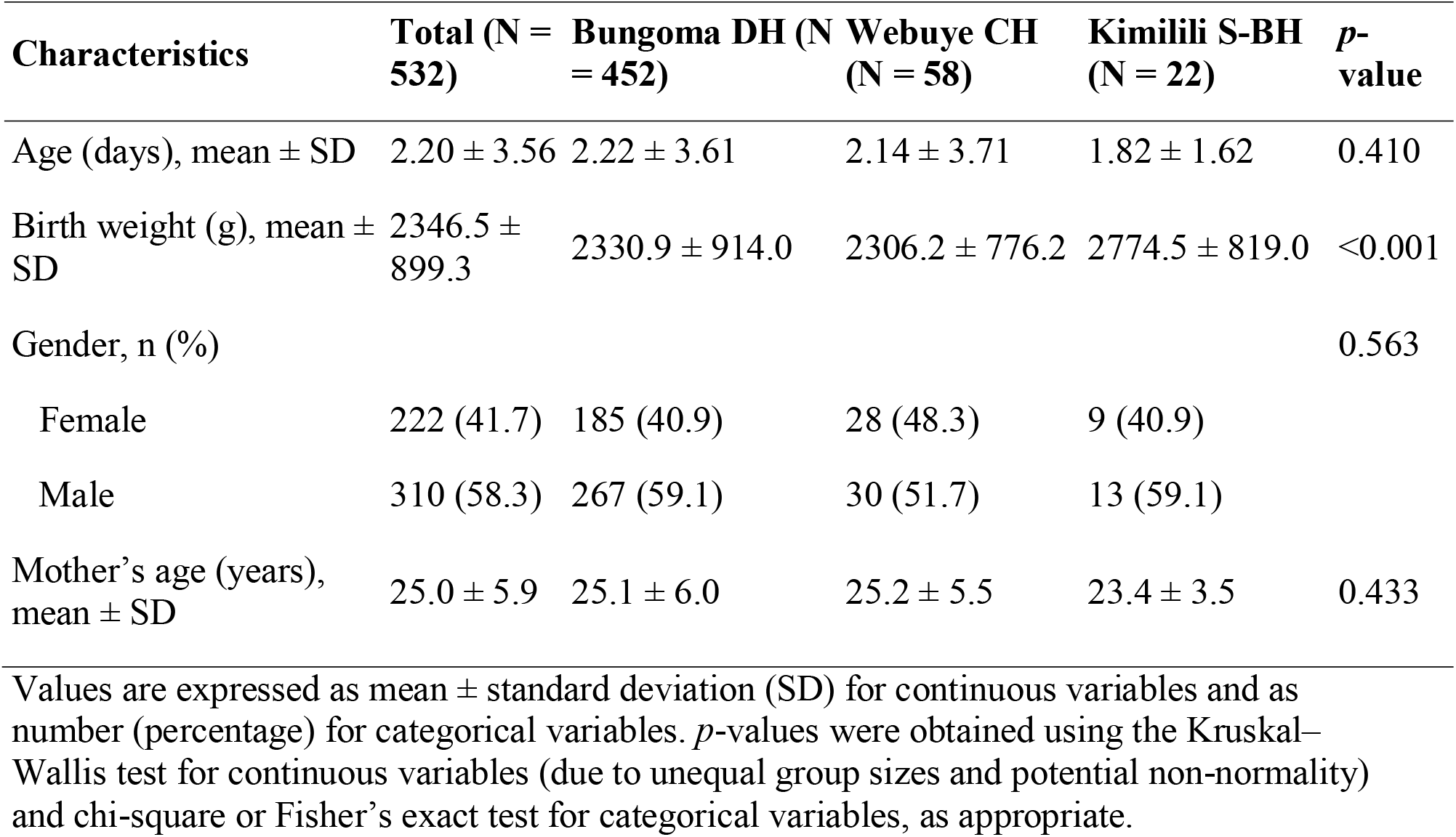
Newborn and maternal characteristics across hospitals.

| Characteristics | Total (N = 532) | Bungoma DH (N = 452) | Webuye CH (N = 58) | Kimilili S-BH (N = 22) | *p*-value |
| --- | --- | --- | --- | --- | --- |
| Age (days), mean ± SD | 2.20 ± 3.56 | 2.22 ± 3.61 | 2.14 ± 3.71 | 1.82 ± 1.62 | 0.410 |
| Birth weight (g), mean ± SD | 2346.5 ± 899.3 | 2330.9 ± 914.0 | 2306.2 ± 776.2 | 2774.5 ± 819.0 | <0.001 |
| Gender, n (%) |  |  |  |  | 0.563 |
| Female | 222 (41.7) | 185 (40.9) | 28 (48.3) | 9 (40.9) |  |
| Male | 310 (58.3) | 267 (59.1) | 30 (51.7) | 13 (59.1) |  |
| Mother's age (years), mean ± SD | 25.0 ± 5.9 | 25.1 ± 6.0 | 25.2 ± 5.5 | 23.4 ± 3.5 | 0.433 |

Values are expressed as mean ± standard deviation (SD) for continuous variables and as number (percentage) for categorical variables. *p*-values were obtained using the Kruskal–Wallis test for continuous variables (due to unequal group sizes and potential non-normality) and chi-square or Fisher's exact test for categorical variables, as appropriate.

**Table 2.**
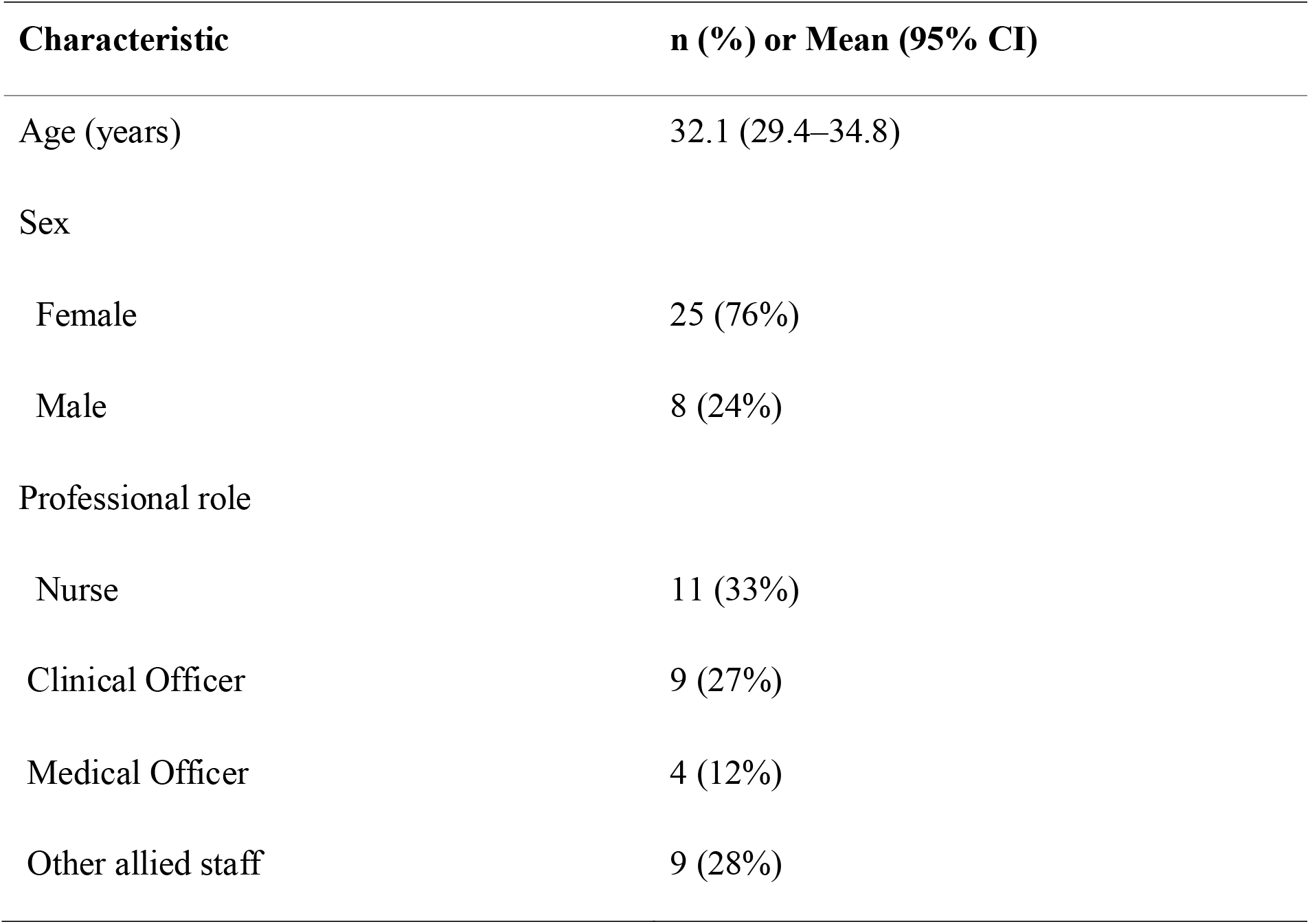
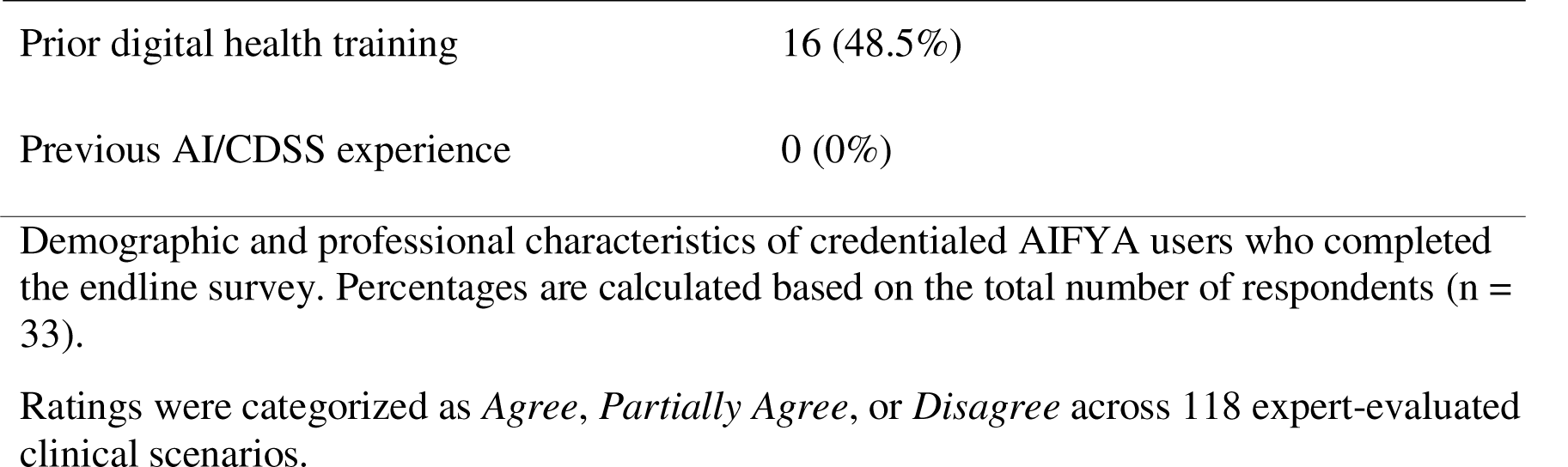
Characteristics of participating healthcare workers.

| Characteristic | n (%) or Mean (95% CI) |
| --- | --- |
| Age (years) | 32.1 (29.4–34.8) |
| Sex |  |
| Female | 25 (76%) |
| Male | 8 (24%) |
| Professional role |  |
| Nurse | 11 (33%) |
| Clinical Officer | 9 (27%) |
| Medical Officer | 4 (12%) |
| Other allied staff | 9 (28%) |
| Prior digital health training | 16 (48.5%) |
| Previous AI/CDSS experience | 0 (0%) |

Demographic and professional characteristics of credentialed AIFYA users who completed the endline survey. Percentages are calculated based on the total number of respondents (n = 33).

### Guideline and citation concordance

Blinded expert evaluation was conducted to assess the correctness of AIFYA-generated neonatal management recommendations against the Kenyan national newborn care protocols. Across 118 clinical scenarios, 75% of the recommendations were rated as correct, 15% as partially correct, and 10% as incorrect (Figure 1). The overall agreement between AIFYA outputs and expert assessments was strong, with a linearly weighted Cohen’s kappa of 0.85 (95% CI: 0.79–0.91). Citation accuracy was also high, with 96% of references correctly cited.

**Figure 1.**
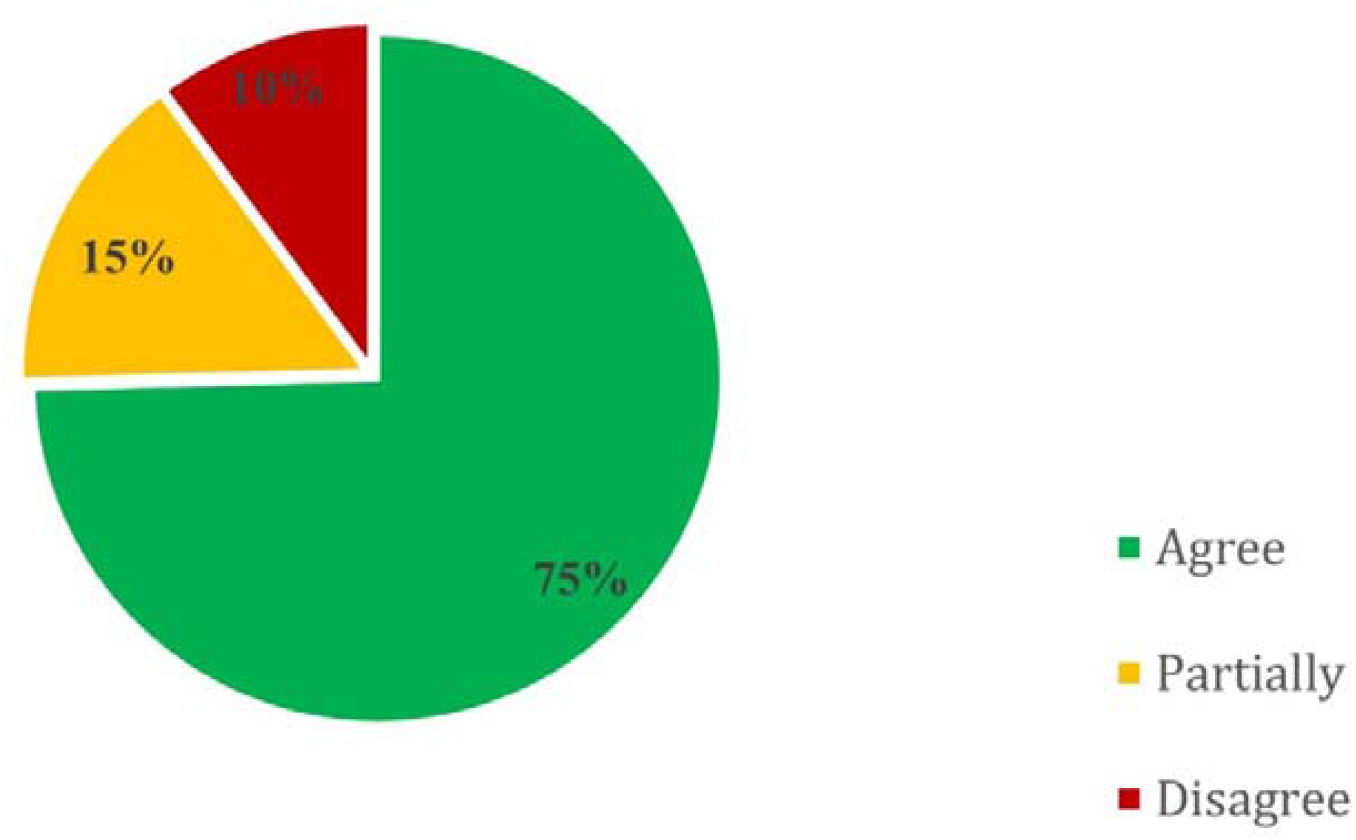
Distribution of AIFYA-generated recommendation ratings. Ratings were categorized as *Agree, Partially Agree*, or *Disagree* across 118 selected expert-evaluated clinical scenarios.

In a focused analysis of 40 complex dosing and fluid management scenarios, experts rated 75% (n = 30) as correct, 15% (n = 6) as partially correct, and 10% (n = 4) as incorrect (Table 3). Most discrepancies arose from weight-based adjustments for extremely preterm infants, and these insights were subsequently used to refine the system’s dosing algorithms and safety guardrails.

**Table 3.** Expert review of AIFYA recommendation correctness and citation accuracy.

| Domain | Correct n (%) | Partially Correct n (%) | Incorrect n (%) | Weighted $\kappa$ (95% CI) |
| --- | --- | --- | --- | --- |
| Overall (n=118 scenarios) | 88 (75%) | 18 (15%) | 12 (10%) | 0.85 (0.79–0.91) |
| Citation correctness | 113 (96%) | – | 5 (4%) | – |
| Focused dosing scenarios (n=40) | 30 (75%) | 6 (15%) | 4 (10%) | – |
| Results of blinded expert evaluation of AIFYA-generated neonatal management recommendations compared to Kenyan national newborn protocols. Agreement statistics are calculated using linearly weighted Cohen’s kappa. |  |  |  |  |

### Process performance and implementation fidelity

Protocol adherence, assessed using a standardized checklist of essential newborn care actions, showed a statistically significant improvement over the study period. The odds of adherence increased by 5% per month (OR = 1.05; 95% CI: 1.02–1.08; p = 0.004), indicating progressive integration of the AIFYA decision support system into routine clinical practice. The median time from patient triage to the first documented clinical decision was 23 minutes (IQR 18–31), and this performance metric remained stable throughout the evaluation period.

The system’s offline-first architecture ensured continuity of service, with 35% of all clinical sessions conducted fully offline due to intermittent internet connectivity. User satisfaction was high, with 92% of healthcare workers (HCWs) agreeing or strongly agreeing that AIFYA was a useful tool in their daily clinical work (Table 4).

**Table 4.** Implementation and process performance outcomes.

| Metric | Result (95% CI or IQR) | Comment |
| --- | --- | --- |
| Total neonatal cases managed via AIFYA | 532 | Across both study sites |
| HCWs trained and credentialed | 50 | Two-day training program |
| Triage-to-decision time (minutes) | 23 (IQR 18–31) | Stable throughout study |
| Sessions completed offline | 35% | Offline-first functionality |
| Change in guideline adherence over time (OR per month) | 1.05 (1.02–1.08), $p=0.004$ | Mixed-effects model clustered by facility |
| User satisfaction (“AIFYA is useful”) | 92% agree or strongly agree | Based on 5-point Likert survey |

Process and implementation metrics summarizing adoption, system performance, and user experience during the 10-month evaluation.

### User perceptions of AI and human oversight

Survey data revealed strong support for a human-governed AI model as summarized in Table 5. A majority of HCWs (79%) rated human oversight of AI-assisted recommendations as “extremely important” or “very important.” The most frequently cited concern regarding AI integration was the potential for clinical over-dependence on the technology (51.5%), followed by concerns about the accuracy of AI-generated recommendations (30.3%) (Table 5).

**Table 5.**
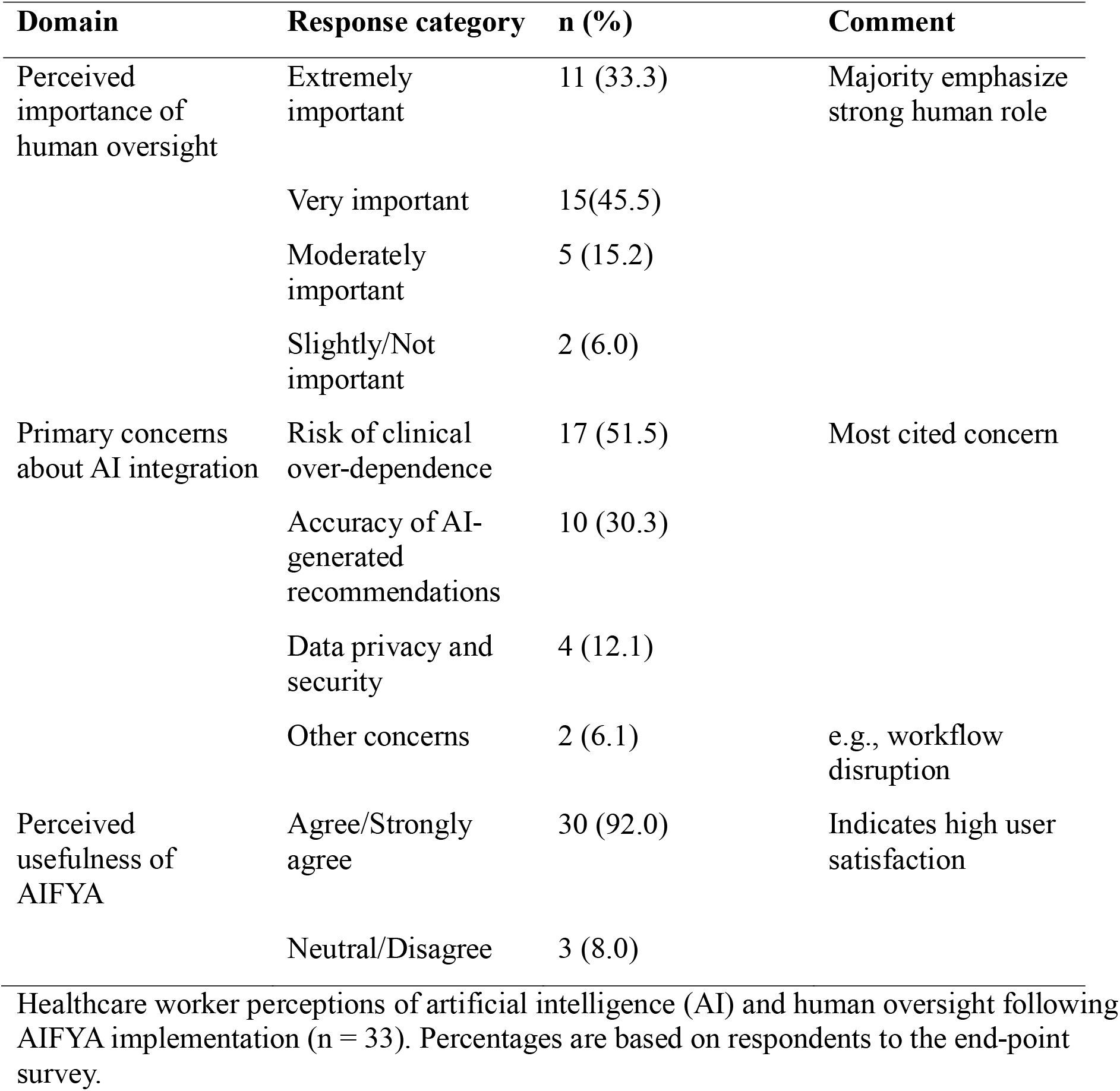
Healthcare worker perceptions of AI use and human oversight in clinical decision-making.

## Discussion

In this pragmatic, early-stage evaluation, we found that a human-supervised, large language model (LLM)–based clinical decision support system (CDSS), carefully aligned with national newborn care guidelines, achieved high adoption, strong clinical concordance, and reliable technical performance when deployed in routine neonatal units in rural Kenya. Together, these findings highlight the feasibility, safety, and acceptability of integrating generative AI tools into frontline clinical workflows in low-resource settings.

First, adoption and reach were substantial, with 550 neonatal cases managed using AIFYA across four facilities and 50 healthcare workers (HCWs) trained and credentialed. The high utilization and satisfaction rates (92% of users agreeing that AIFYA was useful) underscore strong user engagement and confidence in the system. This success was facilitated by a socio-technical design that combined robust offline-first functionality essential in settings with intermittent connectivity with a human-led coaching model. Our findings are consistent with growing literature demonstrating that responsible AI integration, strong human oversight, and context-appropriate design can strengthen clinical practice in resource-constrained environments[10,11].

Second, clinical and technical performance was consistently strong. Blinded expert evaluation revealed 75% correct and 15% partially correct recommendations, with a high overall agreement (κ = 0.85) relative to national newborn protocols. Citation accuracy reached 96%, introducing a novel and transparent measure of “explainable correctness” that enables clinical supervision and fosters user trust. This level of concordance is consistent with emerging evidence showing that well-supervised AI systems can reliably align with established clinical guidelines[12,13]. Notably, focused analysis of dosing and fluid management scenarios revealed that most discrepancies arose from weight-based calculations in extremely preterm infants’ findings that directly informed refinement of the system’s dosing guardrails.

Third, process performance and implementation fidelity demonstrated meaningful improvement in protocol adherence over time. The odds of adherence increased by 5% per month (OR = 1.05; 95% CI: 1.02–1.08; p = 0.004), suggesting that the CDSS supported consistent reinforcement of guideline-based care. The median triage-to-decision time (23 minutes) remained stable throughout the study, indicating that AIFYA integrated smoothly into clinical workflows without introducing delays. Sustaining digital health tools within large and complex health systems requires careful attention to adoption and implementation factors, as documented in previous studies[14–16]. Taken together, these findings highlight that meaningful improvements in clinical practice can be achieved without compromising efficiency or increasing clinician burden.

Finally, user perceptions of AI and human oversight highlight both enthusiasm and caution among frontline clinicians. The majority (79%) emphasized that human oversight of AI-assisted recommendations is “extremely” or “very” important, reflecting a nuanced understanding of AI’s role as a decision-support not decision-making tool. The most frequently cited concerns, including clinical over-dependence (51.5%) and potential inaccuracies (30.3%), reinforce the importance of maintaining a mandatory human-in-the-loop framework. Such concerns mirror well-documented risks of automation bias and error propagation in digital decision-support systems, particularly in resource-constrained settings[10,17,18].

Collectively, these findings provide one of the first real-world examples of an ethically grounded, explainable, and context-adapted AI system deployed in a low-resource clinical setting. By pairing technical innovation with human supervision and local guideline alignment, AIFYA demonstrates a scalable model for safe, responsible, and sustainable AI integration in global health.

### Strengths and limitations

The primary strength of this study lies in its pragmatic design, which evaluated the system under authentic clinical conditions in rural neonatal units. The study’s adherence to the DECIDE-AI reporting framework ensures methodological transparency and reproducibility. Additional strengths include the rigorous, blinded multi-expert review of AI-generated recommendations, providing an objective measure of clinical concordance, and the introduction of “citation correctness” as a novel performance metric for explainability and accountability in LLM-based CDSS.

Several limitations should be acknowledged. As an early-stage evaluation, the study was not powered to assess downstream clinical outcomes such as mortality, morbidity, or length of stay. The observational design limits causal inference, and the presence of study personnel may have introduced a Hawthorne effect, potentially enhancing adherence and performance indicators. Although cases spanned key neonatal conditions, the findings may not be fully generalizable to all neonatal presentations or other healthcare contexts. Additionally, while the offline-first architecture was effective, long-term sustainability and integration within national digital health systems will require further assessment at scale.

## Conclusion

A human-supervised, citation-backed AI decision support system aligned with national newborn care protocols can be deployed safely and effectively within busy, low-resource neonatal units. By transparently anchoring its recommendations to trusted national guidelines, AIFYA strengthens clinical trust, improves adherence to evidence-based care, and demonstrates a responsible framework for integrating large language models into frontline health systems. These findings provide compelling justification for a multi-site, cluster-randomized controlled trial to determine the system’s impact on neonatal morbidity and mortality, and to inform policy-level integration into national digital health strategies.

## Statements

### Data availability statement

The datasets generated and analysed during this study, which supported the development and evaluation of AIFYA, a human-supervised, large language model (LLM)-based clinical decision support system (CDSS) aligned with the Kenya Consolidated Newborn Clinical Guidelines are publicly available. All anonymized datasets can be accessed via Figshare at: http://10.6084/m9.figshare.30836888, under the terms of the Creative Commons Attribution 4.0 International Public License (CC BY 4.0). The source code, including system architecture, implementation details, and technical specifications, is not publicly available due to intellectual property restrictions. Access may be granted by the corresponding author upon reasonable written request (>).

### Ethics statement

This study was approved by the Mount Kenya University Institutional Scientific and Ethics Review Committee (MKU ISERC, ref: MKU/ISERC/3425). Informed consent was secured in writing. Written informed consent was obtained from all participating healthcare workers, and separate written informed consent was obtained from the parents of all newborns whose data were included in the study.

### Author contributions

TK: Methodology, Investigation, Project Administration; GK: Software, Validation, Resources, Writing – Review and Editing; FM: Investigation, Clinical validation, Writing – Review and Editing; PO: Methodology, Formal Analysis (Quantitative), Data Curation, Visualization, Writing – Review and Editing; GM: Data collection, analysis, Review and editing; DK: Methodology, Validation, Writing – Review and Editing SM: Methodology, Formal Analysis (Qualitative), Investigation, Writing – Review and Editing; JG: Conceptualization, Methodology, Supervision, Funding Acquisition, Writing – Original Draft, Writing – Review and Editing. All authors read and approved the final manuscript.

### Funding

This study was supported by the Science for Africa Foundation under grant number GC Africa Round 15: Artificial Intelligence (AI) Seed Grant - GCA/SFA/R15/152 awarded to JG. The funder had no role in the study design, data collection, data analysis, data interpretation, manuscript preparation, or the decision to submit for publication. The views expressed are those of the authors and do not necessarily reflect the positions of the Science for Africa Foundation.

## Acknowledgements

We extend our sincere gratitude to the Bungoma County Ministry of Health, the facility managers, and all healthcare workers across the three participating health facilities for their invaluable time, collaboration, and insights. We also thank the expert physicians who contributed to the validation exercise, as well as the parents and neonates whose clinical data made this research possible.

## Competing interests

The authors declare that they have no competing interests.

## Generative AI statement

The author(s) declared that generative AI was not used in the creation of this manuscript.

Any alternative text (alt text) provided alongside figures in this article has been generated by Frontiers with the support of artificial intelligence and reasonable efforts have been made to ensure accuracy, including review by the authors wherever possible. If you identify any issues, please contact us.

## Supplementary material

## Notes

### Competing Interest Statement

The authors have declared no competing interest.

### Author Declarations

Ethical approval was obtained from the Mount Kenya University Institutional Ethics Review Committee (MKU/IERC/3425), and administrative approval was granted by the Bungoma County Health Management Team (CHMT).

